# Scaling Cues to Freeze Gait: An Augmented Reality Approach to Benchmark Freezing of Gait in Parkinson’s Disease

**DOI:** 10.64898/2026.01.13.25343234

**Authors:** Yara N. Derungs, Charlotte Lang, Carolina I. Conde, William R. Taylor, Mathias Bannwart, Deepak K. Ravi, Chris Easthope Awai

## Abstract

**Background:** Freezing of gait (FOG) affects up to 80% of people with advanced Parkinson’s disease and is difficult to elicit reliably during clinical assessments. Augmented reality (AR) offers potential for standardized FOG provocation by presenting virtual triggers in any environment.

**Objective:** To evaluate whether an AR-based turning task could elicit FOG in a graded, dose-dependent manner and assess user experience with the technology.

**Methods:** Thirteen people with Parkinson’s disease (8 freezers, 5 non-freezers) completed an AR pillar-turning protocol across two cohorts: clinic-based (n=4, all freezers) and laboratory-based (n=9, mixed). Participants performed 360° turns around a virtual pillar presented at three diameters (0.6, 0.4, 0.2 m) using a Microsoft HoloLens 2, manipulating turning radius to vary task difficulty. FOG episodes were video-recorded and independently annotated. Participants completed perception questionnaires and the New Freezing of Gait Questionnaire (NFOG-Q).

**Results:** All clinic freezers exhibited FOG during AR pillar turns, with a clear dose-response relationship: mean episodes increased from 2.3 at 0.6 m to 5.3 at 0.4 m to 8.5 at 0.2 m diameter. No laboratory participants experienced FOG during pillar turns, though one lab freezer froze during return turns. NFOG-Q profiles indicated comparable daily-life FOG severity between clinic and laboratory freezers, suggesting environmental factors drove differential outcomes. Participants reported positive experiences with AR quality, safety, naturalness of movement, and rapid adaptation, though clinic participants reported higher immersion than laboratory participants.

**Conclusions:** AR-based pillar-turning successfully elicited graded FOG in susceptible individuals within a FOG-provoking environment, demonstrating proof-of-concept for scalable virtual trigger paradigms. Effectiveness depends on matching environmental context to individual FOG susceptibility, with implications for standardized clinical FOG assessment.

## Introduction

Parkinson’s disease (PD) is a common neurodegenerative disorder whose global prevalence and burden has more than doubled in recent decades [1], [2]. Alongside the cardinal motor features (bradykinesia, rigidity, tremor, postural instability) [1], progression to advanced PD is often accompanied by worsening gait instability [3] and episodic gait disturbances, most notably freezing of gait (FOG) [4]. FOG is defined as “a brief, episodic absence or marked reduction of forward progression of the feet despite the intention to walk” [4]. It affects around 50% of people with PD (PwPD) in earlier disease stages, rising to ~80% in advanced PD [5], [6], [7]. FOG episodes are heterogeneous in manifestation (e.g. start hesitation, shuffling or trembling in place) and are strongly associated with falls, loss of independence, and reduced quality of life [4], [8], [9], [10], [11]. Currently, despite the significant need, there are no validated therapeutic pathways for FOG [12].

FOG is often difficult to elicit and measure during routine clinical visits [13]. Its paroxysmal and context-dependent nature means that patients show reduced or uncharacteristic freezing behaviour in clinical environments [13], [14], [15]. The mere act of observation can transiently improve gait in PD, likely due to increased attentional focus, reducing the likelihood of capturing FOG in brief clinical exams [16]. Clinicians often rely on patient self-report or questionnaires (e.g. FOG-Q, NFOG-Q), but retrospective recall is prone to bias and brief episodes may be missed [17], [18]. Although various FOG-provocation protocols have been proposed, their use and implementation differ substantially across centers [19]. This variability makes it difficult to compare assessments or establish consistent clinical benchmarks [20]. As a result, there remains no consensus on how to reliably elicit FOG in a controlled setting, despite its importance for both clinical evaluation and mechanistic research [20].

To address these limitations, a growing body of work is focused on identifying and standardizing tasks that reliably provoke FOG [20], [21]. A variety of candidate triggers have been studied – including gait initiation, narrow doorways and passages, sudden obstacles, dual-task interference, and turning in place or while walking [8], [22], [23]. However, their effectiveness in provoking FOG has been inconsistent, with substantial variability reported across studies. A recent systematic review of 128 studies found that turning (especially ~360° turns) was the most effective task for inducing FOG [20]. This aligns with real-life observations, as turning is a frequent and demanding component of daily mobility that requires coordinated whole-body reorientation [24], [25], [26], [27]. Despite this progress, current triggering methods are quite heterogeneous and rely on clinic-specific features, such as obstacle-rich walking paths, or adaptive spatial setups such as doorways that can be made narrower or wider. These solutions still only offer limited control over environmental and cognitive factors, underscoring the need for more flexible and standardized approaches to elicit FOG in realistic yet controlled conditions.

Immersive technologies have recently opened new avenues to investigate FOG triggers in a manner that overcomes many of the limitations of traditional provocation tasks. Virtual reality (VR) enables the creation of safe, repeatable simulations of environments laden with FOG triggers (e.g. virtual doorways, narrow corridors, 360° turns) [28], [29]. VR offers a promising approach to elicit FOG by combining multiple environmental and cognitive triggers in a configurable scenario [28], [29]. Such VR approaches illustrate the potential for standardized FOG elicitation outside of traditional large gait labs. However, fully immersive VR also has practical drawbacks – notably, users are isolated from the real world, and may experience cybersickness, collisions with real world objects, or even be more prone to falls [30].

Augmented reality (AR) offers an exciting alternative for delivering FoG triggers. AR headsets overlay holographic visual stimuli onto the user’s actual surroundings, rather than replacing reality entirely [31]. One advantage of AR over VR is that the patient remains grounded in a real physical environment and peripherally perceives their limbs, which may reduce disorientation and facilitate natural movement. AR devices are able to map the user’s room, enabling the anchoring of digital objects within a user’s current environment. Therefore, they can present context-specific triggers, such as a virtual obstacle appearing at a doorway, while the user still perceives the floor and walls, potentially enhancing safety and realism [32].

The ability of immersive digital technologies to provide scalable, on-demand triggers in any setting, ranging from a clinic hallway to a patient’s home, remains highly appealing [20]. Traditional clinical setups rely on fixed physical configurations (e.g., one doorway width or a single turning cone), which often fail to elicit FOG in patients who report frequent episodes in their daily environments [22]. These static configurations provide only limited challenge and cannot reproduce the subtle variations in spatial or cognitive demand that commonly precipitate FOG in natural settings [22]. In contrast, digital and immersive technologies allow task difficulty to be adjusted dynamically: virtual pathways can be progressively narrowed, turning demands increased, or additional cognitive load introduced. This enables individualized titration of difficulty while preserving standardization across assessments. Notably, such systems do not require large dedicated spaces, enable consistent cue presentation across clinics, and can be deployed relatively inexpensively compared to dedicated infrastructure, such as width-adaptive doorways [33].

Building on these observations, we sought to evaluate an AR-based paradigm for intentionally triggering FOG in a controlled and progressively more challenging manner. In this proof-of-concept study, we developed an AR-based 360° turning task based on the observation that 360° turns are among the most effective FOG triggers [20]. Using a head-mounted AR display, we presented patients with a life-sized virtual pillar (column) in their real environment and instructed them to walk around it, thereby simulating a 360° turn. By adjusting the pillar’s diameter, we could manipulate the turning radius required: from a wide easy turn to increasingly tight turns, which were expected to provoke more frequent FOG episodes. Our paradigm thus provides a single-trigger, scalable FOG challenge that can be deployed even in a small room. We evaluated this AR task in a cohort of PwPD, collecting both quantitative data (FOG occurrences, durations) and qualitative feedback on the user experience.

We hypothesized that (1) an AR-delivered pillar-turning task would successfully elicit FOG episodes in participants with a prior history of FOG (demonstrating the feasibility of AR as a FOG trigger platform), and (2) decreasing the turn radius (smaller pillar) would lead to more frequent or severe FOG events, consistent with prior reports that very tight turns are especially freezing-provoking [20]. In sum, this work introduces and gives preliminary evidence for a novel AR-based approach for graded FOG triggering, thereby adding a (first) puzzle piece to the development of scalable AR trigger paradigms to standardize FOG assessment.

## Methods

### Study Design and Participants

Participants were recruited via convenience sampling from different institutions (clinics, physiotherapists) in Zurich and surrounding areas in two phases. Initially, a small group of people with Parkinson’s disease was recruited to evaluate the functioning of the AR environment in a clinical setting. This was followed by recruitment of an additional cohort that was assessed in a laboratory setting. All participants completed an identical AR-based turning task using a head-mounted display (HMD), with task design, instrumentation, and procedures held constant across cohorts.

Inclusion criteria required participants to be adults with idiopathic PD who were able to walk at least 15 meters independently. Exclusion criteria included other neurological or cognitive comorbidities affecting gait, relevant visual impairments that could interfere with AR display use, and recent major musculoskeletal surgery (within the past 18 months).

The first cohort consisted of four PwPD recruited from the University Hospital Zurich. This clinic-based cohort served as a rapid feasibility test of the AR protocol, chosen primarily due to ease of patient access. Testing took place within the clinic environment in an underground transport corridor. In a clarification of responsibility inquiry (BASEC Nr. Req-2021-00955), the Ethics Committee of the Canton of Zurich determined that this project did not fall within the scope of the Human Research Act and therefore did not require authorization from the ethics committee. The promising results from this initial cohort motivated progression to the second phase with more controlled laboratory-based measurements.

The second cohort consisted of nine PwPD. Testing was conducted in a brightly lit, spacious laboratory at the Laboratory for Movement Biomechanics at ETH Zurich. This protocol was reviewed and approved by the ETH Zurich Ethics Commission (2022-N-156).

Following informed consent and screening, all participants completed demographic and clinical assessments, including the New Freezing of Gait Questionnaire (NFOG-Q). FOG status was determined based on NFOG-Q item 1: freezers were defined by a score of 1 (indicating at least one freezing episode in the previous month) and non-freezers by a score of 0. All participants were tested during their self-reported ON-medication state to approximate everyday mobility (Population details: Table 1).

**Table 1:** Participant characteristics stratified by testing environment and FOG status. Clinic-FOG participants (n=4) were all freezers with longer disease duration and higher DBS prevalence (75%). Lab cohort (n=9) included both freezers (Lab-FOG, n=4) with intermediate disease duration and 50% DBS prevalence, and non-freezers (Lab-NoFOG, n=5) with substantially shorter disease duration and no DBS. FOG manifestation duration was comparable between Clinic-FOG and Lab-FOG groups (3.3 vs 3.6 years), despite marked differences in FOG episodes observed during AR testing. Values presented as mean ± SD.

| | Clinic-FOG ($n=4$) | Lab-FOG ($n=4$) | Lab-NoFOG ($n=5$) |
| --- | --- | --- | --- |
| **Age (mean ± SD)** | $69.0 \pm 9.1$ | $64.0 \pm 7.6$ | $65.6 \pm 13.1$ |
| **Sex (M/F)** | 4/0 | 4/0 | 4/1 |
| **Disease duration (yrs)** | $15.3 \pm 4.0$ | $13.0 \pm 5.3$ | $4.8 \pm 2.2$ |
| **DBS (% with)** | 75% | 50% | 0% |
| **FOG Manifestation (yrs)** | $3.3 \pm 3.3$<br>(range: 0.5 – 8) | $3.6 \pm 4.3$<br>(range: 1 – 10) | – |
| **Total FOG Episodes** | $28.5 \pm 24.6$ | $2.0 \pm 4.0$ | 0 |

### Experimental Conditions

AR stimuli were presented via a Microsoft HoloLens 2 (Microsoft, Redmond, WA). To identify and later annotate FOG episodes, a GoPro® action camera recorded foot motion from an egocentric chest mounting. An overview of the experimental design is given in Figure 1.

**Figure 1:**
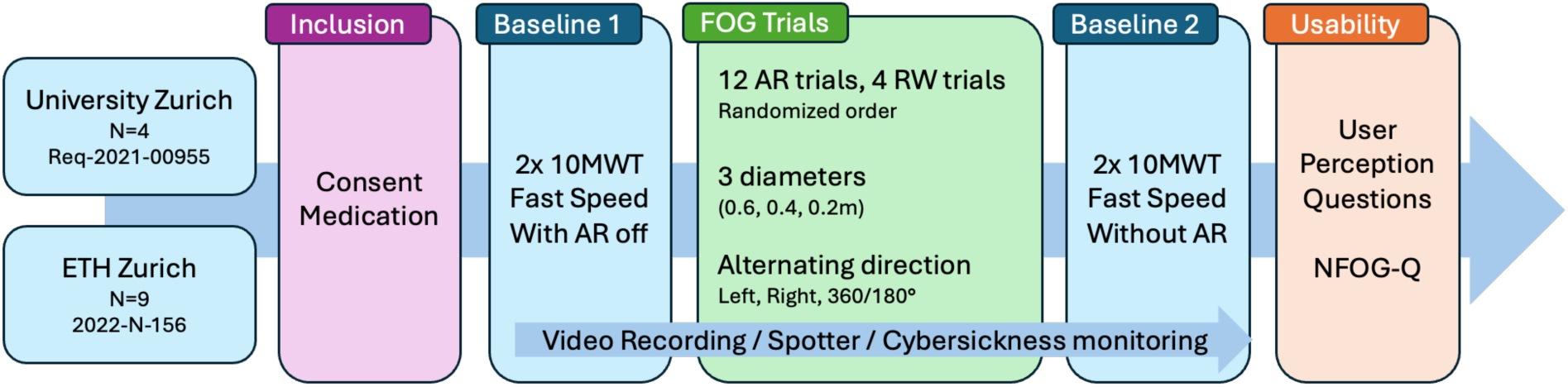
Experimental protocol. Participants were recruited via convenience sampling in two phases: clinic-based feasibility testing (n=4, BASEC Nr. Req-2021-00955) and laboratory-based assessment (n=9, ETH Ethics 2022-N-156). After consent and medication status verification, participants completed baseline walking (2× 10MWT with HoloLens), AR pillar-turning trials (12 trials total: 4 trials per diameter across 3 diameters [0.6, 0.4, 0.2 m] in randomized order; each trial consisted of a 360° walk around the virtual pillar followed by a 180° return turn to starting position; turn direction alternated left/right), real-world turning trials (RW; 4 trials), baseline walking (2× 10MWT without HoloLens), and usability questionnaires (perception and NFOG-Q). Safety and cybersickness monitoring, video recording, and trained spotter supervision were continuous throughout.

For baseline assessment, participants completed two fast-speed 10-m walk tests (10MWT) without turns, prior to the AR block, while wearing the HoloLens 2. In the AR environment, only a virtual cylindrical pillar was displayed, anchored in place. Each trial involved a straight-line approach to the pillar, a full 360° walk around it, and a straight-line departure to a visible finish mark, where participants completed a natural 180° turn to prepare for the next walking trial (Figure 2). No virtual pillar was present during the natural turn; only the finish line marker was visible. To introduce a graded trigger for FOG, the pillar was presented at three diameters - 0.6 m, 0.4 m, and 0.2 m - thereby manipulating the turning radius and increasing task difficulty with smaller diameters. Each participant completed 12 trials in total, with four trials per diameter. Trial order was randomized and turn direction alternated between left and right across trials.

**Figure 2:**
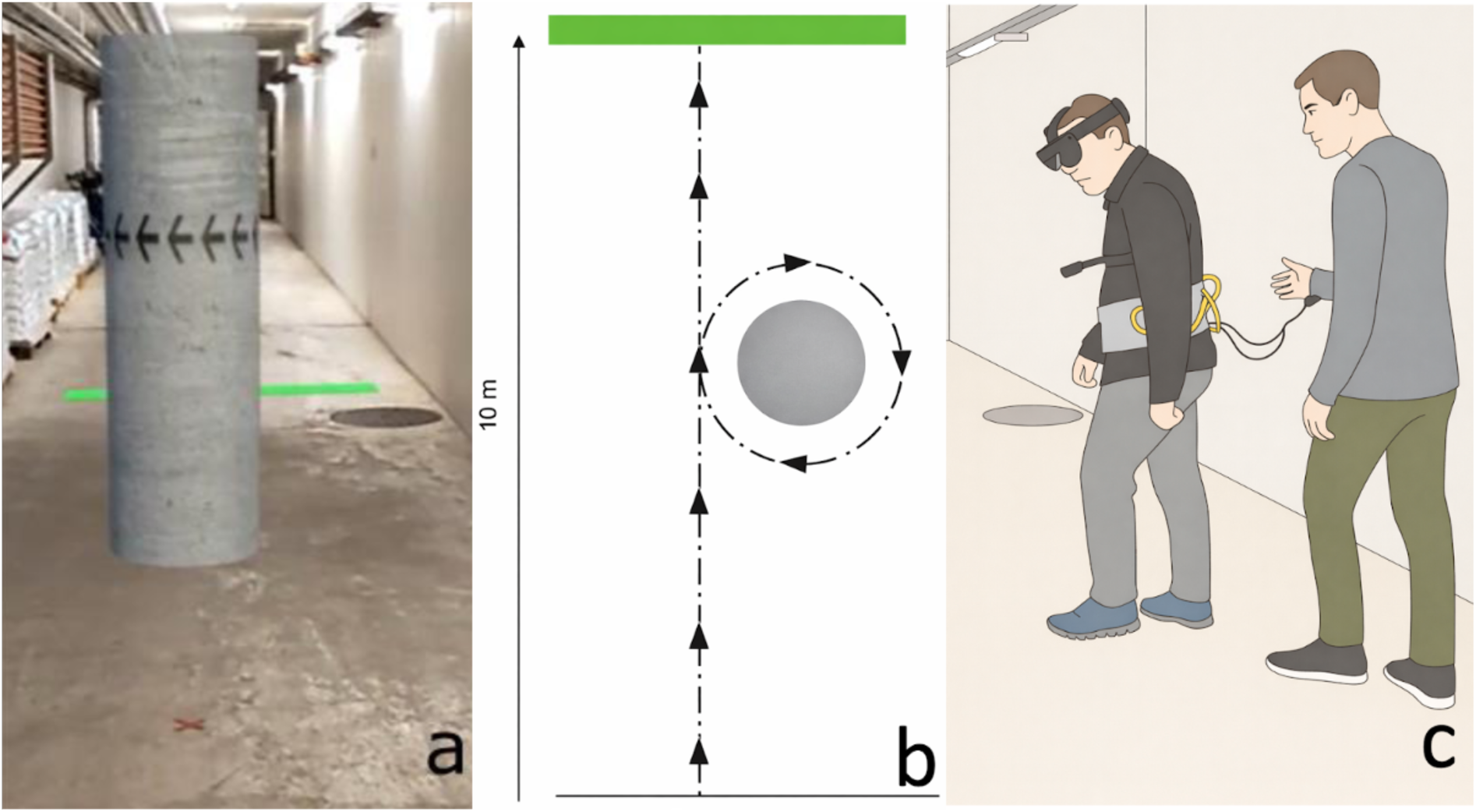
AR pillar-turning task setup and execution. (a) Testing environment in underground transport corridor (clinic cohort) showing physical pillar and green floor markers for start/finish positions. (b) Schematic of AR task structure: participants performed straight-line approach (10 m), complete 360° walk around virtual pillar (shown in gray), and straight-line departure to finish marker (green line). Pillar presented at three diameters (0.6, 0.4, 0.2 m) to manipulate turning radius. (c) Participant wearing Microsoft HoloLens 2 with chest-mounted GoPro camera and safety belt with tethers connected to wrist straps worn by trained spotter.

Following the AR block, participants completed two additional fast-speed 10MWT without the headset, serving as baseline measurements for the real-world (RW) version of the task. In the RW condition, participants walked around a physical floor target at a self-selected turning radius, replicating the same approach–turn–depart structure used in the AR trials. Four RW trials were completed.

Throughout all testing, participants wore a safety belt with tethers, and a trained spotter walked behind them and held these tethers to prevent falls. Sessions were monitored for cybersickness or fatigue through regular verbal check-ins with participants, and testing was paused or discontinued if safety concerns arose. Participants additionally completed usability questionnaires at the end of the session, including a custom perception questionnaire assessing AR experience quality, immersion, safety, and comfort (23 items on a 5-point Likert scale), and the New Freezing of Gait Questionnaire (NFOG-Q) [34] to characterize FOG severity in daily life.

### Video Annotation and Data Processing

Video recordings were segmented by trial. Trials with technical issues or obscured views were excluded and documented. Two independent raters, blinded to condition and diameter, independently annotated FOG episodes using the standard clinical definition: a brief, episodic absence or marked reduction of forward progression despite the intention to walk. The onset and offset of each FOG episode were identified frame-by-frame. Discrepancies exceeding 0.5 seconds between raters were resolved by consensus discussion.

For each trial, we computed the presence or absence of FOG, the number of FOG episodes, the total duration of freezing, and the percentage of time spent frozen.

## Results

### Participant Characteristics

The clinic cohort (ClinicFOG) comprised male participants only (mean age 69.0 ± 9.1 years), all classified as freezers based on NFOG-Q item 1. Mean disease duration was 15.3 ± 4.0 years, with freezing of gait symptoms present for 3.3 ± 3.3 years (range 0.5-8 years).

The laboratory cohort included predominantly male participants (8 male, 1 female), with a mean age of 64.9 ± 10.4 years and a mean disease duration of 8.4 ± 5.6 years. This cohort included both freezers (n=4, Lab-FOG) and non-freezers (n=5, Lab-NoFOG). The Lab-FOG had a mean age of 64.0 ± 7.6 years and mean disease duration of 13.0 ± 5.3 years. The mean duration of FOG manifestation was 3.6 ± 4.3 years (range: 1-10 years). The Lab-NoFOG had a mean age of 65.6 ± 13.1 years and substantially shorter disease duration of 4.8 ± 2.2 years. More population details are available in the supplements (Table S1 – S4).

### FOG Occurrence During AR Pillar-Turning Task

All four clinic participants exhibited FOG during the AR protocol, with a mean of 28.5 ± 24.6 total episodes per participant (range: 4-60). The majority of these episodes occurred during the pillar-turning component of the task. Additional FOG episodes were observed during other phases, such as start hesitation at task initiation.

Focusing on FOG during pillar turns, a dose-response relationship emerged between turning diameter and FOG frequency. As the pillar diameter decreased (tighter turns required), FOG episodes increased progressively: from a mean of 2.3 episodes at 0.6 m diameter, to 5.3 episodes at 0.4 m diameter, and 8.5 episodes at the smallest 0.2 m diameter (Figure 3, Supplements S5).

**Figure 3:**
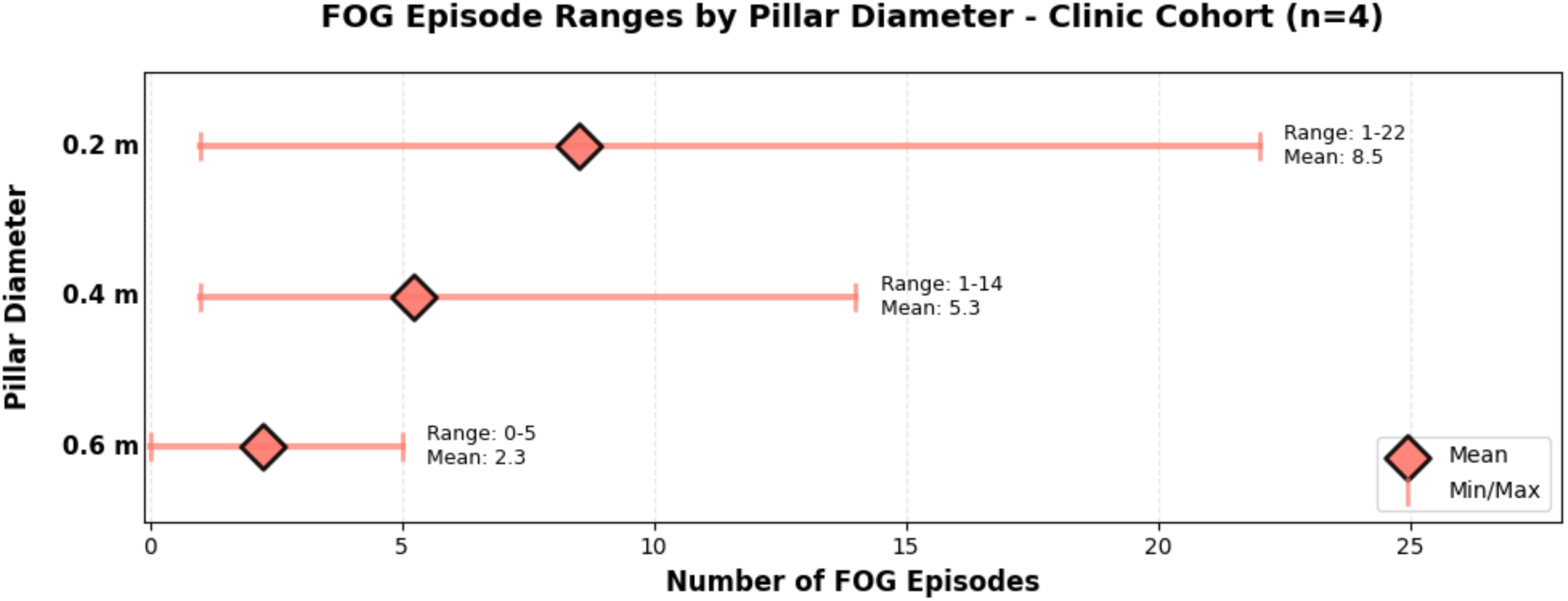
Dose-response relationship between pillar diameter and FOG frequency during pillar turns in clinic cohort (n=4). Clear dose-response relationship observed: as pillar diameter decreased (tighter turns required), FOG frequency increased progressively from 0.6 m diameter (range 0-5, mean 2.3 episodes), to 0.4 m (range 1-14, mean 5.3 episodes), to 0.2 m (range 1-22, mean 8.5 episodes).

This progressive increase with tighter turns was observed in all four participants, even though the individual numbers of FOG episodes varied greatly between participants.

None of the four lab freezers exhibited FOG during the AR pillar-turning task at any radius. However, one freezer experienced 8 FOG episodes during the 180° return turn to the starting position (not during the pillar turns themselves). The remaining three lab freezers showed no FOG during any portion of the protocol.

As expected, none of the five non-freezers exhibited FOG during any trial.

### NFOG-Q Characteristics of Freezers

Among the eight freezers across both cohorts, NFOG-Q responses indicated varying FOG severity in daily life.

Regarding FOG frequency during turning, responses indicated generally high severity: six of eight freezers (75%) reported “very often, more than once per day,” one reported “often, about once per day,” and one reported never experiencing FOG during turns. Episode duration during turning was typically brief to moderate, with most freezers reporting either 2-5 seconds (n=3) or 5-30 seconds (n=2), though one freezer reported episodes as “very long, unable to walk for more than 30 seconds.”

FOG during gait initiation was similarly common, with five of eight freezers (62.5%) reporting “very often, more than once per day.” The impact of FOG on daily activities varied: two freezers rated the impact as severe, two as moderate, two as minimal (“a little”), and two as having no impact at all.

Overall, clinic and laboratory freezers showed similar NFOG-Q profiles in daily life.

### User Experience and Perceptions

Participants generally reported positive experiences with the AR system (Figure 4). Most participants (11 out of 13) agreed or strongly agreed they could perceive the AR environment well. However, two Lab-FOG participants reported difficulty perceiving the AR; one of these was the participant who experienced FOG during the return turns. Nearly all participants (12 out of 13) felt the holograms were well integrated into the real world, and all participants agreed the hologram had a real effect on them. Image quality was rated as good by 11 out of 13 participants, with two neutral responses.

**Figure 4:**
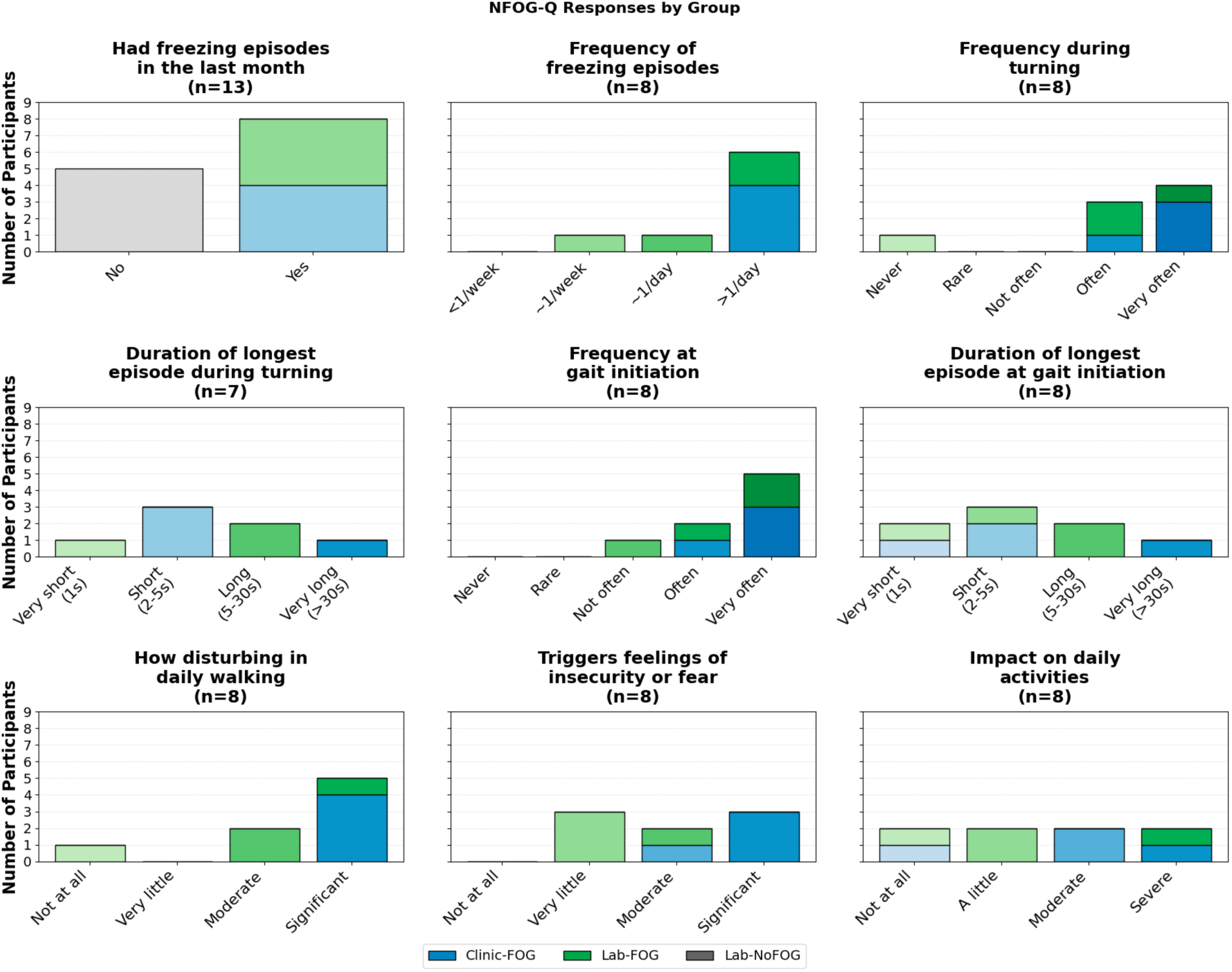
NFOG-Q responses. Bar charts show distribution of participant counts across response categories for nine questions assessing FOG characteristics in daily life. Top row: FOG presence in past month (n=13 including non-freezers), frequency of episodes, and frequency during turning. Middle row: Duration of longest episode during turning (n=7, one freezer did not answer since he does not experience FOG during turning), frequency at gait initiation, and duration at gait initiation. Bottom row: How disturbing FOG is during daily walking, whether it triggers feelings of insecurity or fear, and impact on daily activities. Darker green indicates higher severity/frequency. Clinic-FOG and Lab-FOG groups showed comparable NFOG-Q profiles in daily life despite different FOG outcomes during AR testing, with most reporting very high frequency during turning (75% >1/day) and varied impact on daily activities (25% severe, 25% moderate, 25% minimal, 25% none). Colours represent groups with gradient intensity indicating response level (lighter=disagree, darker=agree).

Movement naturalness was rated highly (12 out of 13), and adaptation to the technology was rapid (12 out of 13 agreed they quickly got used to walking with the HoloLens). Safety perceptions were positive, with 12 out of 13 participants feeling safe walking with the HoloLens, and comfort was rated highly (10 out of 13 agreed the HoloLens was comfortable to wear).

Regarding FOG-specific experiences, most participants did not report increased FOG with the headset. When asked about more gait blockades than usual, 11 out of 13 disagreed, 2 were neutral, and none agreed. Similarly, 12 out of 13 disagreed they experienced longer blockades.

Field of vision showed mixed responses. Two participants (1 Lab-FOG, 1 Lab-NoFOG) reported not being able to perceive everything around them with the HoloLens. Nine participants felt unrestricted in their visual field, 2 were neutral, and 2 felt restricted (the same two who had perception issues). Two Lab-FOG participants noted the field of view was not sufficiently large.

Immersion varied notably by environment. All four Clinic-FOG participants reported forgetting about wearing the HoloLens during the task, whereas all Lab participants either did not forget (7) or were neutral (2). Regarding the sense of forgetting the outside world, 6 out of 13 disagreed (all Lab-NoFOG), while only 3 out of 13 agreed (including one Lab-FOG).

Only one Clinic-FOG participant found the AR experience mentally burdensome. Two Lab-NoFOG participants were disappointed due to higher expectations about AR perception, and one Lab-NoFOG participant reported boredom.

Complete perception questionnaire data are presented in Figure 5 and Supplementary Figure S6.

**Figure 5:**
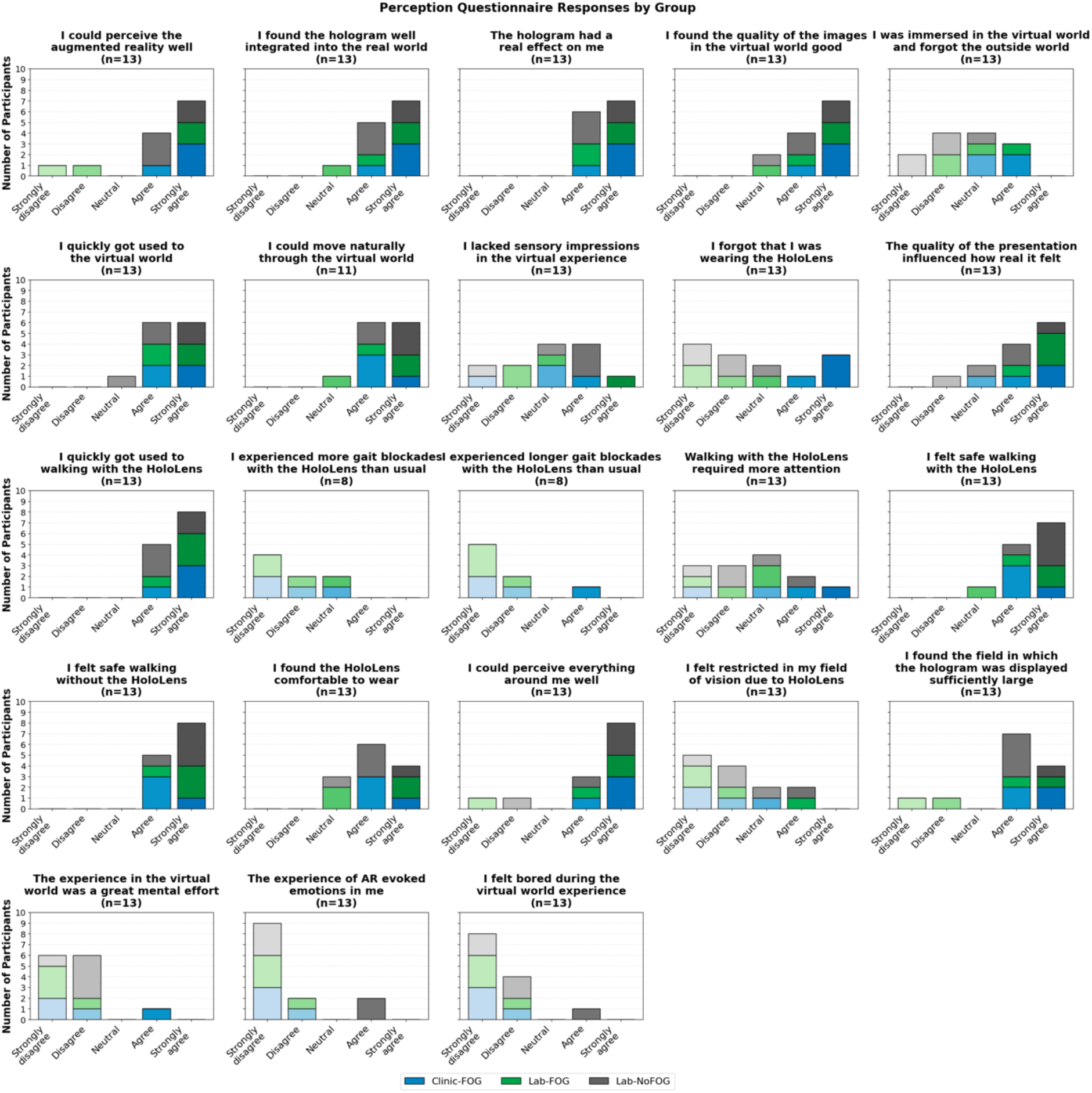
Perception questionnaire responses stratified by testing environment and FOG status (n=13). Stacked bar charts show distribution across 23 items assessing AR experience quality on 5-point Likert scale. Colours represent groups with gradient intensity indicating response level (lighter=disagree, darker=agree). Clinic-FOG participants reported higher immersion (all agreed/strongly agreed to forgetting they were wearing the HoloLens) compared to Lab participants (mostly disagreed). Both cohorts showed positive ratings for AR perception quality, hologram integration, naturalness of movement, rapid adaptation, safety, and comfort. Lab-FOG participants reported more restrictions in field of view compared to other groups. Most participants did not report increased FOG with the headset.

### Safety and Tolerability

No falls or serious adverse events occurred during testing. All participants completed the protocol without cybersickness or substantial discomfort requiring discontinuation. One participant completed 14 trials instead of the planned 20 due to cognitive difficulties with the task, unrelated to safety concerns.

## Discussion

This proof-of-concept study demonstrates that an AR-based turning paradigm can successfully elicit FOG in people with Parkinson’s disease under specific conditions. Our main findings are: (1) An AR pillar-turning task effectively provoked FOG in all freezers tested in the clinic environment, with a dose-response relationship between pillar diameters and FOG frequency; (2) The same protocol did not trigger FOG during pillar turns in the laboratory environment, though one lab freezer experienced FOG during the return turn; (3) Participants across both cohorts reported positive experiences with the AR system, with high ratings for naturalness of movement, safety, and comfort; and (4) The graded turning paradigm (0.6 m → 0.4 m → 0.2 m diameter) provides a scalable approach to quantify FOG severity in a standardized manner.

A key finding of this study is the dose-response relationship between turning radius and freezing in susceptible individuals. Decreasing the virtual pillar diameter required progressively tighter 360° turns and was associated with more frequent FOG episodes in the clinic cohort. This aligns with clinical observations that turning in place and sharper turns, which demand greater axial rotation and postural adjustment, are among the most robust triggers of FOG [20], [24], [25]. Importantly, our AR task reproduces this phenomenon in a controlled, parametric way that requires minimal physical space.

This graded behaviour suggests that turning radius can be used as a scalable trigger to probe an individual’s FOG threshold. A person who only freezes at the smallest radius likely has a higher FOG threshold (milder FOG) than someone who freezes even at wide turns. Such a paradigm could be used to “stress test” the locomotor system and quantify FOG severity in a standardized way across different clinical settings [35]. The minimal space requirements make this approach feasible for small clinics or even home-based assessments, addressing a key limitation of traditional FOG provocation protocols that require large dedicated spaces [33].

The contrast between clinic and laboratory results highlights how environmental context modulates the effectiveness of AR-based FOG triggers. The clinic environment combined several FOG-provoking features, including reduced lighting, a more confined physical space, and a less familiar setting, all of which have been associated with higher FOG risk [23], [36], [37]. In this context, the AR pillar task was sufficient to trigger FOG in all participants. The laboratory, by contrast, provided a wide, well-lit environment with abundant real-world cues that may have stabilized gait.

With the current study design, we cannot completely disentangle environmental from cohort effects. The clinic cohort consisted entirely of established freezers with longer disease duration (15.3 ± 4.0 years) and more advanced therapy (75% with DBS), whereas the lab freezer subgroup had somewhat shorter disease duration (13.0 ± 5.3 years) and lower DBS prevalence (50%). Importantly, despite the different disease profiles, Clinic-FOG and Lab-FOG groups showed comparable FOG profiles in daily life based on NFOG-Q responses: similar proportions reported very frequent turning-related FOG (50% of Clinic-FOG and 50% of Lab-FOG reported >1/day), comparable gait initiation difficulties (75% of Clinic-FOG and 50% of Lab-FOG reported >1/day), and similar duration profiles during turning episodes. This similarity in daily-life FOG severity between Clinic-FOG and Lab-FOG participants supports that the differential FOG outcomes during AR testing were driven primarily by environmental factors rather than baseline FOG severity differences, though the overall longer disease duration in the clinic cohort may have contributed to their susceptibility to environmental triggers.

Despite the technical constraints of early AR hardware, including restricted field of view and limited brightness in well-lit environments [31], [38], [39], participants generally reported positive experiences with the system. The majority felt safe walking with the HoloLens, could move naturally through the virtual world, and adapted quickly to the technology. Importantly, most participants did not experience increased FOG frequency or duration with the headset compared to their usual experience. This high acceptability suggests that AR-based paradigms could be feasibly deployed in clinical practice and research settings.

However, some participants, particularly in the bright laboratory, noted that the holographic pillar was sometimes difficult to see against the background, consistent with known limitations of optical see-through AR displays. Notably, one of the two Lab-FOG participants who reported difficulty perceiving the AR and found the field of view insufficient was the only lab participant who experienced FOG during the experiment (during return turns). This suggests that inadequate AR visibility may have reduced the trigger’s salience for this individual, potentially explaining why FOG occurred only during the non-cued 180° return turn rather than during the AR pillar turns themselves.

Immersion levels also varied by environment. Two Clinic-FOG participants reported forgetting they were in a room wearing the HoloLens, with the other two taking a neutral position. In contrast, only one Lab participant (Lab-NoFOG) reported this level of immersion, while two Lab participants were neutral (one Lab-FOG, one Lab-NoFOG) and the remaining six disagreed. This difference in subjective immersion aligns with the objective FOG outcomes: the dim, confined clinic environment not only provided more FOG-provoking physical conditions but also enhanced psychological engagement with the virtual task, whereas the bright, spacious laboratory may have made the AR elements less salient relative to the rich real-world visual environment.

Most prior AR studies in Parkinson’s disease have focused on cueing to alleviate FOG rather than provoking it. Some work suggests that continuously visible AR floor cues can shorten or prevent FOG episodes and are well received by patients [31], [32], [40]. However, Janssen et al. reported that AR turning targets did not improve FOG during in-place turning and were inferior to rhythmic auditory cues [41]. These mixed results likely reflect differences in cue design, task demands, and device characteristics.

Our study extends this literature by demonstrating that AR can be used not only for cueing but also for systematically provoking FOG with a scalable turning paradigm. This dual capability, encompassing both the triggering and potential alleviation of FOG, positions AR as a versatile platform for FOG research and clinical assessment. The key insight is that AR effectiveness depends on matching the task design, environmental context, and participant characteristics to the intended application.

The successful elicitation of FOG in the clinic cohort, combined with the dose-response relationship across turning radii, supports the feasibility of AR-based FOG assessment in clinical settings. The paradigm’s minimal space requirements and standardized presentation overcome key limitations of traditional provocation protocols, which often require large, dedicated spaces and vary substantially in implementation across centers [17], [18]. For research applications, the ability to systematically vary turning difficulty while maintaining identical visual and cognitive demands opens new possibilities for studying FOG mechanisms. The paradigm could be extended to incorporate additional triggers (e.g., doorways, obstacles, dual tasks) or combined with neuroimaging or electrophysiology to probe the neural correlates of FOG under controlled conditions.

This proof-of-concept study has several limitations that should be acknowledged. First, the small sample size (n=13) and unequal group sizes limit statistical power and generalizability. The confounding of environmental factors with disease severity between cohorts, with the clinic group having longer disease duration and higher DBS prevalence than lab freezers, prevents clear attribution of the differential FOG outcomes to environmental versus individual factors. Future controlled studies should test the same individuals across different environmental conditions to isolate these effects.

Second, the technical constraints of early AR hardware (HoloLens 2) limited display brightness, contrast, and field of view, particularly in well-lit environments [31], [38], [39]. These limitations reduced AR visibility for some participants and may have diminished trigger salience in the laboratory setting. The rapid advancement of AR/XR technology suggests these hardware limitations will be substantially reduced in next-generation devices.

Third, the convenience sampling approach and differences in recruitment sources between cohorts introduce potential selection bias. The clinic cohort represented more advanced disease with established FOG, whereas the lab cohort was more heterogeneous. A more balanced design with severity-matched groups across environments would strengthen causal inferences.

Finally, our outcome measures focused on FOG occurrence and frequency but did not include detailed kinematic or electrophysiological measures that could provide mechanistic insights into why the paradigm succeeded in one environment but not the other. Future work incorporating such measures would enhance understanding of the mechanisms underlying environment-dependent FOG triggering.

## Conclusion

This proof-of-concept study demonstrates that an AR-based pillar-turning paradigm can successfully elicit FOG in people with Parkinson’s disease in a graded, dose-dependent manner, but effectiveness is strongly modulated by environmental context and individual disease characteristics. The dose-response relationship between turning radius and FOG frequency in the clinic cohort validates the core concept of using parametric AR tasks to probe FOG thresholds in a standardized way. High user acceptance and positive safety outcomes support the feasibility of deploying such paradigms in clinical and research settings.

While the failure to elicit FOG during AR pillar turns in the laboratory environment—likely reflecting both less FOG-provoking environmental conditions and differences in disease severity—highlights important constraints, it does not diminish the value of the approach. Rather, it underscores that AR-based FOG triggers, like traditional physical triggers, must be calibrated to individual susceptibility and contextual factors. The minimal space requirements, standardized delivery, and scalable difficulty adjustment offered by AR paradigms address critical limitations of traditional FOG provocation protocols.

As AR hardware continues to improve, with wider fields of view, higher brightness, and better integration of virtual elements into real-world scenes, the effectiveness and applicability of such paradigms are likely to increase substantially. Future work should focus on testing severity-matched individuals across controlled environmental manipulations, extending the paradigm to incorporate multiple FOG triggers, and validating sensitivity to therapeutic interventions. The promising results from this initial study support continued development of AR-based approaches as complementary tools for FOG assessment, mechanism research, and potentially intervention in Parkinson’s disease.

## Data Availability

The data that support the findings of this study are available on request from the corresponding author (CEA). The data are not publicly available due to their containing information that could compromise the privacy of research participants. Video data is not available due to ethical and privacy restrictions concerning the identifiable nature of the supporting data.

## Code Availability

The codebase for the augmented reality triggering tool is freely available for use at OSF under an MIT license. The code is provided unsupported, with no warranties, and as-is: https://osf.io/fyc3h/overview?view_only=869879c11c7a47f5bdcc8ed0fc3adcc0

## Generative AI-use Statement

Claude Sonnet 4.5 was used to improve sentence construction.

## References

[1] R. Balestrino and A. H. V. Schapira, ‘Parkinson disease’, Eur. J. Neurol., vol. 27, no. 1, pp. 27–42, 2020, doi: 10.1111/ene.14108.

[2] E. R. Dorsey et al., ‘Global, regional, and national burden of Parkinson’s disease, 1990–2016: a systematic analysis for the Global Burden of Disease Study 2016’, Lancet Neurol., vol. 17, no. 11, pp. 939–953, Nov. 2018, doi: 10.1016/S1474-4422(18)30295-3.

[3] C. Lang et al., ‘Foot placement coordination is impaired in people with Parkinson’s disease’, J. Neuroengineering Rehabil., Dec. 2025, doi: 10.1186/s12984-025-01830-6.

[4] J. G. Nutt, B. R. Bloem, N. Giladi, M. Hallett, F. B. Horak, and A. Nieuwboer, ‘Freezing of gait: moving forward on a mysterious clinical phenomenon’, Lancet Neurol., vol. 10, no. 8, pp. 734–744, Aug. 2011, doi: 10.1016/S1474-4422(11)70143-0.

[5] D. M. Tan, J. L. McGinley, M. E. Danoudis, R. Iansek, and M. E. Morris, ‘Freezing of Gait and Activity Limitations in People With Parkinson’s Disease’, Arch. Phys. Med. Rehabil., vol. 92, no. 7, pp. 1159–1165, July 2011, doi: 10.1016/j.apmr.2011.02.003.

[6] M. A. Hely, W. G. J. Reid, M. A. Adena, G. M. Halliday, and J. G. L. Morris, ‘The Sydney multicenter study of Parkinson’s disease: The inevitability of dementia at 20 years’, Mov. Disord., vol. 23, no. 6, pp. 837–844, 2008, doi: 10.1002/mds.21956.

[7] S. Perez-Lloret et al., ‘Prevalence, Determinants, and Effect on Quality of Life of Freezing of Gait in Parkinson Disease’, JAMA Neurol., vol. 71, no. 7, pp. 884–890, July 2014, doi: 10.1001/jamaneurol.2014.753.

[8] M. Falla, G. Cossu, and A. Di Fonzo, ‘Freezing of gait: overview on etiology, treatment, and future directions’, Neurol. Sci., vol. 43, no. 3, pp. 1627–1639, Mar. 2022, doi: 10.1007/s10072-021-05796-w.

[9] Y. Okuma, A. L. Silva de Lima, J. Fukae, B. R. Bloem, and A. H. Snijders, ‘A prospective study of falls in relation to freezing of gait and response fluctuations in Parkinson’s disease’, Parkinsonism Relat. Disord., vol. 46, pp. 30–35, Jan. 2018, doi: 10.1016/j.parkreldis.2017.10.013.

[10] C. C. Walton et al., ‘The major impact of freezing of gait on quality of life in Parkinson’s disease’, J. Neurol., vol. 262, no. 1, pp. 108–115, Jan. 2015, doi: 10.1007/s00415-014-7524-3.

[11] B. R. Bloem, J. M. Hausdorff, J. E. Visser, and N. Giladi, ‘Falls and freezing of gait in Parkinson’s disease: A review of two interconnected, episodic phenomena’, Mov. Disord., vol. 19, no. 8, pp. 871–884, 2004, doi: 10.1002/mds.20115.

[12] C. K. Cui and S. J. G. Lewis, ‘Future Therapeutic Strategies for Freezing of Gait in Parkinson’s Disease’, Front. Hum. Neurosci., vol. 15, p. 741918, Nov. 2021, doi: 10.3389/fnhum.2021.741918.

[13] C. Barthel, E. Mallia, B. Debû, B. R. Bloem, and M. U. Ferraye, ‘The Practicalities of Assessing Freezing of Gait’, J. Park. Dis., vol. 6, no. 4, pp. 667–674, doi: 10.3233/JPD-160927.

[14] J. Spildooren, S. Vercruysse, K. Desloovere, W. Vandenberghe, E. Kerckhofs, and A. Nieuwboer, ‘Freezing of gait in Parkinson’s disease: The impact of dual-tasking and turning’, Mov. Disord., vol. 25, no. 15, pp. 2563–2570, 2010, doi: 10.1002/mds.23327.

[15] M. Sawada, K. Wada-Isoe, R. Hanajima, and K. Nakashima, ‘Clinical features of freezing of gait in Parkinson’s disease patients’, Brain Behav., vol. 9, no. 4, p. e01244, 2019, doi: 10.1002/brb3.1244.

[16] J. F. Hafer, R. Vitali, R. Gurchiek, C. Curtze, P. Shull, and S. M. Cain, ‘Challenges and advances in the use of wearable sensors for lower extremity biomechanics’, J. Biomech., vol. 157, p. 111714, Aug. 2023, doi: 10.1016/j.jbiomech.2023.111714.

[17] N. Giladi and A. Nieuwboer, ‘Understanding and treating freezing of gait in parkinsonism, proposed working definition, and setting the stage’, Mov. Disord., vol. 23, no. S2, pp. S423–S425, 2008, doi: 10.1002/mds.21927.

[18] F. Hulzinga et al., ‘The New Freezing of Gait Questionnaire: Unsuitable as an Outcome in Clinical Trials?’, Mov. Disord. Clin. Pract., vol. 7, no. 2, pp. 199–205, 2020, doi: 10.1002/mdc3.12893.

[19] A. Cucca et al., ‘Freezing of Gait in Parkinson’s Disease: From Pathophysiology to Emerging Therapies’, Neurodegener. Dis. Manag., vol. 6, no. 5, pp. 431–446, Oct. 2016, doi: 10.2217/nmt-2016-0018.

[20] C. I. Conde, C. Lang, C. R. Baumann, C. A. Easthope, W. R. Taylor, and D. K. Ravi, ‘Triggers for freezing of gait in individuals with Parkinson’s disease: a systematic review’, Front. Neurol., vol. 14, p. 1326300, Dec. 2023, doi: 10.3389/fneur.2023.1326300.

[21] J. O’Day, J. Syrkin-Nikolau, C. Anidi, L. Kidzinski, S. Delp, and H. Bronte-Stewart, ‘The turning and barrier course reveals gait parameters for detecting freezing of gait and measuring the efficacy of deep brain stimulation’, PLoS ONE, vol. 15, no. 4, p. e0231984, Apr. 2020, doi: 10.1371/journal.pone.0231984.

[22] M. Mancini, B. R. Bloem, F. B. Horak, S. J. G. Lewis, A. Nieuwboer, and J. Nonnekes, ‘Clinical and methodological challenges for assessing freezing of gait: Future perspectives’, Mov. Disord., vol. 34, no. 6, pp. 783–790, 2019, doi: 10.1002/mds.27709.

[23] S. R. Lord et al., ‘Freezing of Gait in People with Parkinson’s Disease: Nature, Occurrence, and Risk Factors’, J. Park. Dis., vol. 10, no. 2, pp. 631–640, Apr. 2020, doi: 10.3233/JPD-191813.

[24] B. C. Glaister, G. C. Bernatz, G. K. Klute, and M. S. Orendurff, ‘Video task analysis of turning during activities of daily living’, Gait Posture, vol. 25, no. 2, pp. 289–294, Feb. 2007, doi: 10.1016/j.gaitpost.2006.04.003.

[25] F. Huxham, R. Baker, M. E. Morris, and R. Iansek, ‘Head and trunk rotation during walking turns in Parkinson’s disease’, Mov. Disord., vol. 23, no. 10, pp. 1391–1397, 2008, doi: 10.1002/mds.21943.

[26] P. Crenna et al., ‘The association between impaired turning and normal straight walking in Parkinson’s disease’, Gait Posture, vol. 26, no. 2, pp. 172–178, July 2007, doi: 10.1016/j.gaitpost.2007.04.010.

[27] M. Mancini, A. Weiss, T. Herman, and J. M. Hausdorff, ‘Turn Around Freezing: Community-Living Turning Behavior in People with Parkinson’s Disease’, Front. Neurol., vol. 9, Jan. 2018, doi: 10.3389/fneur.2018.00018.

[28] L. I. Gómez-Jordana, J. Stafford, C. (Lieke) E. Peper, and C. M. Craig, ‘Crossing Virtual Doors: A New Method to Study Gait Impairments and Freezing of Gait in Parkinson’s Disease’, Park. Dis., vol. 2018, no. 1, p. 2957427, 2018, doi: 10.1155/2018/2957427.

[29] T. Siragy et al., ‘VReeze: an open-source virtual reality for the examination of freezing of gait in Parkinson’s disease – a study design of a crossover repeated measures study for validation’, BMJ Open, vol. 15, no. 11, p. e106489, Nov. 2025, doi: 10.1136/bmjopen-2025-106489.

[30] M. Tukur et al., ‘Panoramic imaging in immersive extended reality: a scoping review of technologies, applications, perceptual studies, and user experience challenges’, *Front*. Virtual Real., vol. 6, Sept. 2025, doi: 10.3389/frvir.2025.1622605.

[31] E. M. Hoogendoorn, D. J. Geerse, J. Helsloot, B. Coolen, J. F. Stins, and M. Roerdink, ‘A larger augmented-reality field of view improves interaction with nearby holographic objects’, PLOS ONE, vol. 19, no. 10, p. e0311804, Oct. 2024, doi: 10.1371/journal.pone.0311804.

[32] B. Baugher, N. Szewczyk, and J. Liao, ‘Augmented reality cueing for freezing of gait: Reviewing an emerging therapy’, Parkinsonism Relat. Disord., vol. 116, p. 105834, Nov. 2023, doi: 10.1016/j.parkreldis.2023.105834.

[33] B. Bluett, E. Bayram, and I. Litvan, ‘The Virtual Reality of Parkinson’s Disease Freezing of Gait: A Systematic Review’, Parkinsonism Relat. Disord., vol. 61, pp. 26–33, Apr. 2019, doi: 10.1016/j.parkreldis.2018.11.013.

[34] A. Nieuwboer et al., ‘Reliability of the new freezing of gait questionnaire: Agreement between patients with Parkinson’s disease and their carers’, Gait Posture, vol. 30, no. 4, pp. 459–463, Nov. 2009, doi: 10.1016/j.gaitpost.2009.07.108.

[35] A. Nieuwboer and N. Giladi, ‘Characterizing freezing of gait in Parkinson’s disease: Models of an episodic phenomenon’, Mov. Disord., vol. 28, no. 11, pp. 1509–1519, 2013, doi: 10.1002/mds.25683.

[36] K. A. E. Martens, C. G. Ellard, and Q. J. Almeida, ‘Does Anxiety Cause Freezing of Gait in Parkinson’s Disease?’, PLOS ONE, vol. 9, no. 9, p. e106561, Sept. 2014, doi: 10.1371/journal.pone.0106561.

[37] M. Ishii and K. Okuyama, ‘Characteristics associated with freezing of gait in actual daily living in Parkinson’s disease’, J. Phys. Ther. Sci., vol. 29, no. 12, pp. 2151–2156, Dec. 2017, doi: 10.1589/jpts.29.2151.

[38] A. Erickson, K. Kim, G. Bruder, and G. F. Welch, ‘Exploring the Limitations of Environment Lighting on Optical See-Through Head-Mounted Displays’, in Symposium on Spatial User Interaction, Virtual Event Canada: ACM, Oct. 2020, pp. 1–8. doi: 10.1145/3385959.3418445.

[39] A. Samini, K. L. Palmerius, and P. Ljung, ‘A Review of Current, Complete Augmented Reality Solutions’, in 2021 International Conference on Cyberworlds (CW), Caen, France: IEEE, Sept. 2021, pp. 49–56. doi: 10.1109/CW52790.2021.00015.

[40] D. J. Geerse, B. Coolen, J. J. van Hilten, and M. Roerdink, ‘Holocue: A Wearable Holographic Cueing Application for Alleviating Freezing of Gait in Parkinson’s Disease’, Front. Neurol., vol. 12, Jan. 2022, doi: 10.3389/fneur.2021.628388.

[41] S. Janssen et al., ‘The Effects of Augmented Reality Visual Cues on Turning in Place in Parkinson’s Disease Patients With Freezing of Gait’, Front. Neurol., vol. 11, Mar. 2020, doi: 10.3389/fneur.2020.00185.

