## Supplementary for "Scaling Cues to Freeze Gait: An Augmented Reality Approach to Benchmark Freezing of Gait in Parkinson’s Disease"

### Appendix

#### Clinic

***Table S1:*** *Individual participant characteristics and FOG episode counts for clinic cohort (n=4). All participants were male freezers with mean age 69.0±9.1 years, disease duration 15.3±4.0 years, and FOG manifestation 3.3±3.3 years. Three had deep brain stimulation (DBS). All were tested in ON-medication state (P3 off dopaminergic medication due to DBS). NFOG-Q scores indicated established FOG in daily life (Part 1=1 for all; Part 2 range: 14-17; Part 3 range: 6-9). Total FOG episodes during testing ranged from 4 to 60 (mean 28.5±24.6), with progressive increase across pillar diameters: 0.6 m diameter (0-5 episodes), 0.4 m (1-14 episodes), 0.2 m (1-22 episodes). P4 completed only 14 trials instead of 20 due to cognitive difficulties with task comprehension. Follow-up (1-2 days post-testing) confirmed no adverse events.*

| | **Participant 1** | **Participant 2** | **Participant 3** | **Participant 4** |
|---|---|---|---|---|
| **sex** | male | male | male | male |
| **Time since disease onset (years before assessment)** | 11 years | 17 years | 13 years | 20 years |
| **Time since FOG onset (years before assessment)** | 2 years | 2.5 years | 6 months | 8 years |
| **medication status** | ON | ON | OFF since DBS | ON |
| **DBS** | No | Yes | Yes | Yes |
| **Clinical assessment** | Freezing of Gait Q<br>Part 1 = 1<br>Part 2 = 15<br>Part 3 = 9 | Freezing of Gait Q<br>Part 1 = 1<br>Part 2 = 14<br>Part 3 = 7 | Freezing of Gait Q<br>Part 1 = 1<br>Part 2 = 15<br>Part 3 = 6 | Freezing of Gait Q<br>Part 1 = 1<br>Part 2 = 17<br>Part 3 = 8 |
| **Turning** | AR first<br>RW second | RW first<br>AR second | AR first<br>RW second | RW first<br>AR second |
| **follow up** | 2 days after<br>no pain or other bad consequences from participating | 1 day after<br>no pain, no exhaustion | 1 day after<br>no pain, no muscle pains, no other consequences | |
| | | | | patient had difficulties to perform walking task for cognitive reasons<br>decided to perform only 6 gait trials in AR condition |

| | | | | |
|---|---|---|---|---|
| | | | | → only 14 gait trials instead of 20 |
| **Total FOG** | 15 | 35 | 60 | 4 |
| **FOG during gait trial** | 5 | 27 | 39 | 3 |
| **FOG during turn** | 4 | 26 | 39 | 3 |
| **start hesitation** | 1 | 1 | 0 | 0 |
| **FOG AR 0.6 m** | 1 | 5 | 3 | 0 |
| **FOG AR 0.4 m** | 1 | 5 | 14 | 1 |
| **FOG AR 0.2 m** | 2 | 9 | 22 | 1 |

### Lab

***Table S2:*** *Individual participant characteristics and FOG episode counts for laboratory cohort (n=9, P5-P13). Participants included 8 males and 1 female with mean age 64.9±10.4 years and disease duration 8.4±5.6 years. Four participants were freezers and five were non-freezers. All tested in ON-medication state. Only P9 experienced FOG during testing (8 episodes total: 6 during gait trials, 5 during 180° return turns, 1 start hesitation). No FOG occurred during AR pillar turns at any diameter (0.6 m, 0.4 m, 0.2 m) for any laboratory participant.*

| | **Participant 5** | **Participant 6** | **Participant 7** | **Participant 8** | **Participant 9** | **Participant 10** | **Participant 11** | **Participant 12** | **Participant 13** |
|---|---|---|---|---|---|---|---|---|---|
| **sex** | M | M | M | M | M | F | M | M | M |
| **Time since disease onset (years before assessment)** | 5 years | 3 years | 2.5 years | 8 years | 20 years | 5.5 years | 14 years | 10 years | 8 years |
| **Time since FOG onset (years before assessment)** | - | - | - | - | 10 years | - | 2 years | 1.5 years | 1 years |
| **medication status** | ON | ON | ON | ON | ON | ON | ON | ON | ON |
| **DBS** | No | No | No | No | No | No | Yes | Yes | No |

| | | | | | | | | | |
|---|---|---|---|---|---|---|---|---|---|
| **Total FOG** | - | - | - | - | 8 | - | - | - | - |
| **FOG during gait trial** | - | - | - | - | 6 | - | - | - | - |
| **FOG during turn** | - | - | - | - | 5 | - | - | - | - |
| **start hesitation** | - | - | - | - | 1 | - | - | - | - |
| **FOG AR 0.6 m** | - | - | - | - | - | - | - | - | - |
| **FOG AR 0.4 m** | - | - | - | - | - | - | - | - | - |
| **FOG AR 0.2 m** | - | - | - | - | - | - | - | - | - |

#### Population comparisons

***Table S3:*** *Comparison of clinic cohort (n=4) and laboratory cohort (n=9) characteristics and FOG outcomes. Clinic cohort consisted entirely of freezers with longer disease duration (15.3±4.0 years) and higher DBS prevalence (75%). Laboratory cohort was more heterogeneous. Total FOG episodes during testing were substantially higher in clinic cohort (28.5±24.6) compared to laboratory cohort, where only one participant (P9) experienced FOG episodes. FOG manifestation duration shown for clinic cohort only (laboratory cohort included both freezers and non-freezers).*

| | **Clinic Cohort (n=4)** | **Lab Cohort (n=9)** |
|---|---|---|
| **Age (mean ± SD)** | 69.0 ± 9.09 | 64.9 ± 10.4 |
| **Sex (M/F)** | 4/0 | 8/1 |
| **Disease duration (yrs)** | 15.3 ± 4.0 | 8.4 ± 5.6 |
| **DBS (% with)** | 75% | |
| **FOG Manifestation (yrs)** | 3.3 ± 3.3<br>(range: 0.5 - 8) | |
| **Freezers (% total)** | 100% | 44.4% |
| **Total FOG Episodes** | 28.5 ± 24.6 | 0.6 ± 2.2 |

***Table S4:*** *Comparison of participant characteristics by cohort and FOG status. Clinic-FOG participants (n=4) were all freezers with longer disease duration (15.3±4.0 years) and higher DBS prevalence (75%). Lab-FOG participants (n=4) had comparable FOG manifestation duration (3.6±4.3 years) and intermediate disease duration (13.0±5.3 years) with 50% DBS prevalence. Lab-NoFOG participants (n=5) had substantially shorter disease duration (4.8±2.2 years) and no DBS. Lab-NoFOG participants experienced no FOG as expected.*

| | **Clinic-FOG (n=4)** | **Lab-FOG (n=4)** | **Lab-NoFOG (n=5)** |
|---|---|---|---|
| **Age (mean ± SD)** | 69.0 ± 9.1 | 64.0 ± 7.6 | 65.6 ± 13.1 |
| **Sex (M/F)** | 4/0 | 4/0 | 4/1 |
| **Disease duration (yrs)** | 15.3 ± 4.0 | 13.0 ± 5.3 | 4.8 ± 2.2 |

| | | | |
|---|---|---|---|
| **DBS (% with)** | 75% | 50% | 0% |
| **FOG Manifestation (yrs)** | 3.3 ± 3.3 (range: 0.5 – 8) | 3.6 ± 4.3 (range: 1 – 10) | – |
| **Freezers (% total)** | 100% | 100% | 0% |
| **Total FOG Episodes** | 28.5 ± 24.6 | 2.0 ± 4.0 | 0 |

***Table S5:*** *FOG episodes during AR pillar-turning task by participant and pillar diameter. Clinic cohort (P1-P4) showed progressive increase in FOG frequency as diameter decreased from 0.6 m to 0.2 m, with total episodes ranging from 4 to 60 per participant (mean 28.5±24.6). Laboratory cohort (P5-P13) showed no FOG during pillar turns at any diameter. P9's total of 8 episodes occurred exclusively during 180° return turns to starting position (not during pillar turns). P9, P11, P12, P13 were classified as freezers based on NFOG-Q item 1=1 but did not experience FOG during the AR pillar-turning protocol. P5-P8, P10 were non-freezers.*

| Participant | FOG AR 0.6 m | FOG AR 0.4 m | FOG AR 0.2 m | Overall FOG Episodes |
|---|---|---|---|---|
| **P1 (Clinic)** | 1 | 1 | 2 | 15 |
| **P2 (Clinic)** | 5 | 5 | 9 | 35 |
| **P3 (Clinic)** | 3 | 14 | 22 | 60 |
| **P4 (Clinic)** | 0 | 1 | 1 | 4 |
| **P5 (Lab)** | 0 | 0 | 0 | 0 |
| **P6 (Lab)** | 0 | 0 | 0 | 0 |
| **P7 (Lab)** | 0 | 0 | 0 | 0 |
| **P8 (Lab)** | 0 | 0 | 0 | 0 |
| **P9 (Lab)** | 0 | 0 | 0 | 8 |
| **P10 (Lab)** | 0 | 0 | 0 | 0 |
| **P11 (Lab)** | 0 | 0 | 0 | 0 |
| **P12 (Lab)** | 0 | 0 | 0 | 0 |
| **P13 (Lab)** | 0 | 0 | 0 | 0 |

#### Detailed Perception Responses

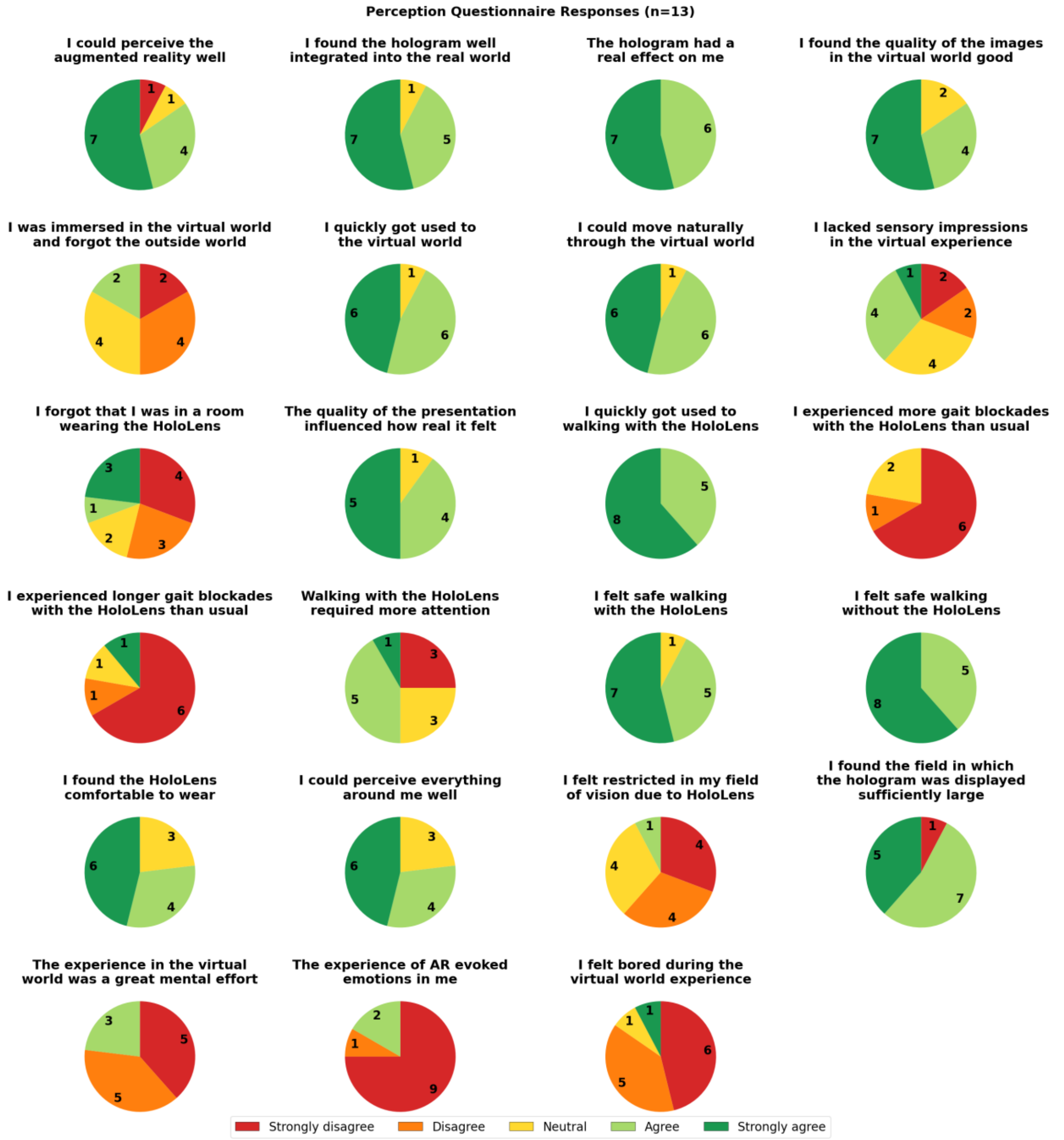


***Figure S6:*** *User perception questionnaire responses (n=13). Pie charts showing distribution across 23 items assessing AR experience on 5-point Likert scale (red=strongly disagree to dark green=strongly agree). Numbers indicate participant counts. Overall positive ratings: AR perception (11/13), hologram integration (12/13), real effect (13/13), movement naturalness (12/13), rapid adaptation (12/13), and safety (12/13 with HoloLens, 13/13 without). Most did not report increased FOG with headset (11/13 denied more blockades, 12/13 denied longer blockades). Environmental differences emerged: Clinic-FOG participants reported higher immersion (forgetting they wore HoloLens), while two Lab-FOG participants (including P8 who experienced FOG during return turns) reported difficulty perceiving AR and insufficient field of view.*
